# Glial activity load on PET (GALP) reveals persistent ‘smoldering’ inflammation in MS despite disease modifying treatment: [F-18]PBR06 study

**DOI:** 10.1101/2023.10.06.23295721

**Authors:** Tarun Singhal, Steven Cicero, Eero Rissanen, John Hunter Ficke, Preksha Kukreja, Steven Vaquerano, Bonnie Glanz, Shipra Dubey, William Sticka, Kyle Seaver, Marie Kijewski, Alexis M. Callen, Renxin Chu, Kelsey Carter, David Silbersweig, Tanuja Chitnis, Rohit Bakshi, Howard L Weiner

## Abstract

**Introduction:** Cortical grey (CoGM) and white matter (WM) microglial activation (MA) is involved in the pathogenesis of multiple sclerosis (MS). [F-18]PBR06 positron emission tomography (PET) targeting 18kilodalton-translocator protein (TSPO) can detect abnormal MA in MS.

**Aims and Objectives:** The goal of this study is to determine the effect of disease modifying treatment (DMT) efficacy on modulating the extent and clinical and radiological correlates of MA in MS patients.

**Methods:** Thirty [F-18]PBR06 PET scans were performed in 22 MS patients (13 RR, 9 SP, mean age 46±14 years, 15 females, median EDSS 3.5, mean T25FW 7.2±4.6s) and 8 healthy controls (HC). Individualized z-score maps of brain parenchymal MA were generated by voxel-by-voxel comparison between each subject’s PET SUVR images and a HC dataset. Logarithmically transformed ‘Glial activity load on PET’ scores (calculated as the sum of voxel-by-voxel z-scores ≥4 in CoGM and WM regions), ‘lnGALP’, were compared between MS subjects on DMT with high efficacy (HT; including rituximab, ocrelizumab, natalizumab and fingolimod, n=13) versus those on no or lower efficacy treatment (LT; including glatiramer acetate and interferons), and correlated with clinical measures and cortical thickness (measured using Freesurfer). p<0.05 was considered statistically significant.

**Results:** CoGM and WM lnGALP scores were higher in MS vs. HCs (10.0±1.5 vs. 7.5±1.5 and 9.8±1.5 vs. 6.6±2.4, both p<0.01) and were inversely correlated with cortical thickness across groups (r=-0.44 and - 0.48, both p<0.05, n=30). In HT-MS group, CoGM and WM lnGALP was significantly lower as compared to LT-MS group (9.1±1.0 vs. 11.3±1.1 and 9.1±1.3 vs. 10.8±1.4, p=0.000075 and 0.006) but remained abnormally higher than in HC group (p=0.006 and 0.02, respectively). Within HT-MS patients, CoGM lnGALP scores were higher in SP vs. RR subgroups (p=0.008), correlated positively with EDSS, T25FW, fatigue scores and serum GFAP levels (r=0.65,0.79, 0.75 and 0.67, all p<0.05), and inversely with cortical thickness (r=-0.66, p=0.01).

**Conclusions:** High-efficacy DMTs decrease, but do not normalize, CoGM and WM MA in MS patients. Such “residual” MA in CoGM is associated with clinical disability, symptom severity and cortical degeneration. Individualized mapping of TSPO-PET using [F-18]PBR06 can potentially serve as an imaging biomarker for evaluating emerging therapies targeting MA in MS patients who are worsening despite high-efficacy DMTs.

## INTRODUCTION

Positron Emission Tomography (PET) imaging using radioligands that target the 18kDa translocator protein (TSPO), located on the outer mitochondrial membrane, has been used to study innate immune activation in multiple sclerosis and various other brain diseases.^1^ Pathological studies have shown that TSPO-PET signal in MS largely originates from CD68+ microglia/macrophages with some contribution from astrocytes, and that TSPO-PET reports on glial density in MS.^2, 3^ ‘Smoldering’ inflammation is a recently popularized term used to describe widespread innate immune activation that is present beyond focal inflammatory lesions in MS patients.^4^ This persistent smoldering inflammation has been considered as a critical component of disease progression but is not readily assessed in MS patients.^4^

[^18^F]PBR06, a phenoxy-aryl-acetamide derivative, is a second generation TSPO ligand which has been validated as a marker of innate immune activation in several preclinical models and has been evaluated in human studies involving MS patients.^5–12^ Our initial studies have revealed that [F-18]PBR06-PET signal is increased in progressive MS as compared to relapsing MS^5^, is linked with fatigue and depressive symptoms^6^, and can demonstrate changes prospectively, following treatment with disease modifying treatment (DMT) in MS patients^13^. We have also shown that in a head-to-head comparison with [C-11]PBR28, [F-18]PBR06-PET measures had a lower coefficient of variation and correlated more strongly with clinical measures in MS subjects.^12^

[F-18]PBR06 has higher affinity (*K _i_* = 0.3 ± 0.08 nM) for its target as compared to [C-11]PK11195(*Ki* = 3.48 ± 1.26 nM), high brain uptake, and a high proportion of specific to nonspecific binding provide high sensitivity to detect small changes in TSPOs in brain.^7, 9, 14^ Metabolic stability of [^18^F]PBR06 relative to the longer half-life of the [^18^F]PBR06 allows the extended acquisition time to measure TSPO as a biomarker of innate immune activation in the brain. Longer half-life of [F-18]PBR06 enables widespread use of this ligand for clinical trials and potential clinical use in future, without the need of an on-site cyclotron. Moreover, routine arterial sampling and metabolite analyses are not clinically feasible^15^ and generally speaking, many assumptions made in tracer kinetic modeling may not be valid and generate parameters that themselves require further validation by other independent measurements, which are of biochemical, clinical and/or pathological significance.^15–17^ Further, it has been stated that mere fitting of a model to kinetic data does not prove the validity of a model.^17^ Hence, our aim is to study a novel, non-invasive, clinically feasible, individualized approach using [F-18]PBR06-PET for identifying widespread smoldering inflammation and its relationship with clinical disability, brain volumetric changes (specifically, cortical atrophy) and serum biomarkers in MS patients.

## METHODS

### Subjects

Thirty [F-18]PBR06-PET scans were performed in 22 MS subjects and 8 healthy controls. Of the 22 MS subjects, thirteen patients were on high-efficacy DMT (HT) (9 women; mean age ± SD, 43 ± 13 years [range, 23–65 years] median Expanded Disability Status Scale (EDSS) score = 4.0 [range, 1.0–7.5]); timed 25-ft walk (T25FW) = 7.47 ± 4.61 seconds (range, 3.40–19.40 seconds)), and 9 patients were on low-efficacy DMT or no DMT (LT) (7 women; mean age ± SD, 50 ± 14 years [range, 28-69 years]; median EDSS score = 3.0 [range, 1.5-6.5]; T25FW = 6.77 ± 4.41 seconds (range, 3.91-17.26 seconds). 8 healthy controls (HC) (3 women; mean age ± SD, 44 ± 17 years [range, 25-69 years]) were also prospectively recruited for this study. MS subjects in the HT group were on rituximab, natalizumab and fingolimod versus those in the LT group were on no DMT or on platform therapies including glatiramer acetate and interferons. Subject characteristics are summarized in Table 1. The study was approved by the Institutional Review Board and Radioactive Drug Research Committee of our hospital; written informed consent was obtained from all participants. The ClinicalTrials.gov registration number for this study was NCT02649985.

**Table 1.** Characteristics Summary.

|  | HT (N=13) | LT (N=9) | HC (N=8) | p Value<br>(HT vs LT) | p Value<br>(HT vs HC) | p Value<br>(LT vs HC) |
| --- | --- | --- | --- | --- | --- | --- |
| **Age (years)** | 43 ± 13 | 50 ± 14 | 44 ± 17 |  |  |  |
| **Sex** | 9 F, 4 M | 7 F, 2 M | 3 F, 5 M |  |  |  |
| **TSPO Binding Affinity** | 8 HAB, 5 MAB | 5 HAB, 4 MAB | 4 HAB, 4 MAB |  |  |  |
| **MS Group** | 6 PMS, 7 RMS | 3 PMS, 6 RMS |  |  |  |  |
| **Median EDSS** | 4.0 | 3.0 |  |  |  |  |
| **T25FW** | 7.47 ± 4.61 | 6.77 ± 4.41 |  |  |  |  |
| **MFIS** | 35 ± 19 | 24 ± 19 |  |  |  |  |

Abbreviations: HT= High-efficacy Treatment; LT= Low-efficacy Treatment; HC= Healthy Control; TSPO= translocator protein; HAB= High Affinity Binder; MAB= Medium Affinity Binder; PMS= Progressive MS; RMS= Relapsing MS; EDSS= Expanded Disability Status Scale; T25FW= Timed 25-Foot Walk; MFIS= Modified Fatigue Impact Score.
Data are mean ± SD, unless otherwise specified.

**Table 2.** Detailed Characteristics.

| Number | Sex | Age Range (y) | TSPO Binding Affinity | MS Group | DMT | EDSS | T25FW | MFIS |
| --- | --- | --- | --- | --- | --- | --- | --- | --- |
| 1 | M | 51-55 | MAB | PMS | Rituximab | 6.5 | 6.17 | 15 |
| 2 | M | 51-55 | HAB | PMS | Rituximab | 6.0 | 8.50 | 37 |
| 3 | F | 56-60 | MAB | PMS | Glatiramer acetate | 6.0 | 12.10 | 54 |
| 4 | F | 56-60 | HAB | PMS | NONE | 6.5 | 17.26 | 71 |
| 5 | F | 46-50 | HAB | PMS | Rituximab | 4.5 | 6.76 | 62 |
| 6 | F | 56-60 | HAB | PMS | Rituximab | 7.5 | 19.40 | 30 |
| 7 | F | 56-60 | MAB | PMS | Rituximab | 6.0 | 7.05 | 66 |
| 8 | F | 61-65 | HAB | PMS | Rituximab | 6.0 | 16.08 | 48 |
| 9 | M | 51-55 | HAB | PMS | Interferon beta-1a | 2.0 | 4.01 | 24 |
| 10 | F | 26-30 | HAB | RMS | NONE | 3.5 | 5.20 | 19 |
| 11 | F | 36-40 | HAB | RMS | Natalizumab | 4.0 | 6.10 | 64 |
| 12 | M | 36-40 | HAB | RMS | Rituximab | 3.0 | 5.00 | 33 |
| 13 | F | 31-35 | MAB | RMS | Fingolimod | 1.0 | 3.40 | 16 |
| 14 | M | 31-35 | MAB | RMS | Fingolimod | 1.0 | 4.00 | 15 |
| 15 | F | 21-25 | HAB | RMS | Natalizumab | 1.5 | 4.80 | 6 |
| 16 | F | 41-45 | HAB | RMS | Fingolimod | 1.5 | 4.99 | 21 |
| 17 | F | 26-30 | MAB | RMS | Fingolimod | 2.0 | 4.83 | 37 |
| 18 | F | 66-70 | HAB | RMS | NONE | 1.5 | 4.50 | 9 |
| 19 | F | 41-45 | HAB | RMS | NONE | 2.0 | 4.75 | 67 |
| 20 | F | 31-35 | MAB | RMS | Glatiramer acetate | 3.0 | 3.91 | 29 |
| 21 | M | 46-50 | MAB | RMS | NONE | 3.5 | 4.55 | 19 |
| 22 | F | 56-60 | MAB | RMS | NONE | 3.0 | 4.69 | 15 |
| 23 | M | 21-25 | MAB | HC |  |  |  |  |
| 24 | F | 41-45 | HAB | HC |  |  |  |  |
| 25 | M | 56-60 | MAB | HC |  |  |  |  |
| 26 | F | 21-25 | HAB | HC |  |  |  |  |
| 27 | F | 31-35 | MAB | HC |  |  |  |  |
| 28 | M | 31-35 | HAB | HC |  |  |  |  |
| 29 | M | 66-70 | MAB | HC |  |  |  |  |
| 30 | M | 61-65 | HAB | HC |  |  |  |  |

Abbreviations: TSPO= translocator protein; HAB= High Affinity Binder; MAB= Medium Affinity Binder; PMS= Progressive MS; RMS= Relapsing MS; HC= Healthy Control; DMT= Disease Modifying Treatment; EDSS= Expanded Disability Status Scale; T25FW= Timed 25-Foot Walk; MFIS= Modified Fatigue Impact Score.

**Table 3a.**
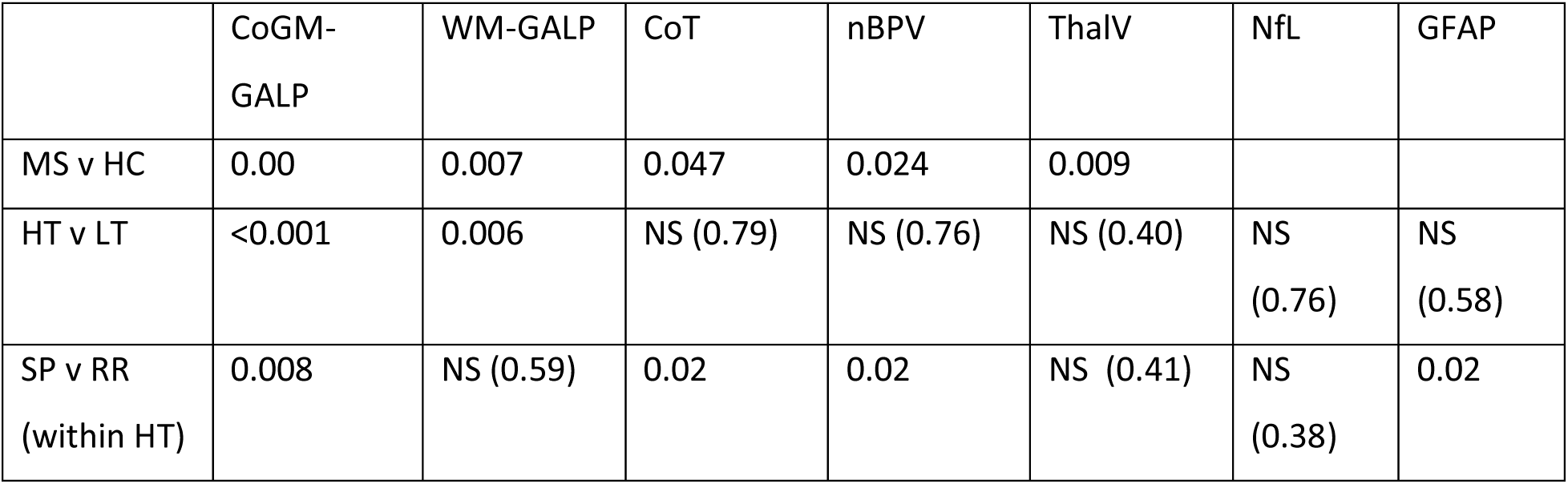

|  | CoGM-GALP | WM-GALP | CoT | nBPV | ThalV | NfL | GFAP |
| --- | --- | --- | --- | --- | --- | --- | --- |
| MS v HC | 0.00 | 0.007 | 0.047 | 0.024 | 0.009 |  |  |
| HT v LT | <0.001 | 0.006 | NS (0.79) | NS (0.76) | NS (0.40) | NS<br>(0.76) | NS<br>(0.58) |
| SP v RR<br>(within HT) | 0.008 | NS (0.59) | 0.02 | 0.02 | NS (0.41) | NS<br>(0.38) | 0.02 |

**Table 3b.** Correlation coefficients and p values for comparisons of clinical measures with PET, MRI and serum biomarkers in HT group.

|  | EDSS | T25FW | MFIS |
| --- | --- | --- | --- |
| CoGM-InGALP | 0.645 [0.017]* | 0.786 [0.001]** | 0.75 [0.003]** |
| WM-InGALP | 0.452 [0.121] | 0.401 [0.174] | 0.39 [0.19] |
| CoT | -0.579 [0.038]* | -0.720 [0.006]** | -0.21 [0.47] |
| nBPV | -0.826 [0.001]** | -0.736 [0.004]** | 0.29 [0.94] |
| Th-V | -0.490 [0.089] | -0.555 [0.049]* | -0.247 [0.595] |
| NfL | 0.49 [0.448] | 0.677 [0.03]* | 0.52 [0.12] |
| GFAP | 0.67 [0.03]* | 0.806 [0.005]** | 0.311 [0.38] |

\* equals $p < 0.05$ ; \*\* equals $p < 0.01$

### Genotyping

Blood samples drawn on the initial screening visit were used to obtain genomic DNA and assess polymorphism within the TSPO gene on chromosome 22q13.2, using a Taqman assay. High-(HAB) and medium-affinity binders (MAB) were included in this study, whereas low-affinity binders were excluded.^18^ Of the 13 HT subjects, 8 were HABs, and 5 were MABs. Of the 9 LT subjects, 5 were HABs and 4 were MABs. Of the 8 HC subjects, 4 were HABs and 4 were MABs.

### Production of Radiopharmaceuticals

18F-PBR06 was produced in the PET radiochemistry facility at our hospital according to standardized procedures.^7, 9, 14^ These were purified by high-pressure liquid chromatography and sterilized by membrane filtration using a 0.22-μm membrane filter. The final product was dispensed in an isotonic solution that was sterile and pyrogen-free and ready for intravenous (IV) administration. The radiochemical purity of the radiopharmaceuticals was determined using high-pressure liquid chromatography. The organic solvents were determined using gas chromatography. The radiochemical purity of each radiopharmaceutical was greater than 95%.

### MRI Acquisition and Analysis

All subjects underwent brain MRI scans on the same scanner (Siemens 3 T Skyra, Siemens Healthineers, Erlangen, Germany) using the same high-resolution acquisition protocol. Whole-brain images were acquired with a 2D T1-weighted spin echo axial series. In addition, patients underwent a 3D magnetization-prepared rapid gradient-echo sequence (MPRAGE, voxel size 1 mm^3^). Only noncontrast MRI scans were obtained; IV gadolinium contrast was not administered as part of this study. Whole brain and thalamic normalized volumes and cortical thickness were measured based on our previously described methods using the MPRAGE sequence.^5, 19–21^

Serum glial fibrillary acid protein (GFAP) and neurofilament light chain (NfL) measurement Serum GFAP and NfL were measured according to our previously described methods.^22^

### PET Acquisition and Analysis

Radiotracers were injected as a bolus for PET scanning using an IV catheter inserted into an upper extremity vein; images were acquired in a list acquisition mode using a high-resolution, whole-body PET/CT scanner. The mean injected doses of 18F-PBR06 were 2.51 ± 0.53 mCi (range, 1.39–3.56 mCi). The mean time interval between MRI and 18F-PBR06 PET scans was 28 ± 42 days (range, 0-182 days). Dynamic images were acquired over 120 minutes and summed 18F-PBR06 PET images were derived based on PET data acquired between 60 and 90 minutes after tracer injection and were then coregistered to each individual patient’s T1 MRI (the spin-echo or MPRAGE) scans using the PMOD 3.8/3.9 platform (PMOD Technologies, Zurich, Switzerland; www.pmod.com). This standardized algorithm involved coregistration of the T1-weighted series and PET images of each individual with the Automated Anatomical Template.^5,6^ Individualized parametric 3-dimensional z-score maps of brain parenchymal MA were generated by voxel-by-voxel statistical comparison between each subject’s globally normalized PET images and a HC dataset.^8^ Logarithmically transformed ‘Glial activity load on PET’ (GALP) scores (calculated as the sum of voxel-by-voxel z-scores thresholded at ≥4 in CoGM and WM regions), ‘lnGALP’, were compared between MS subjects on HT versus LT, and correlated with clinical measures, serum biomarkers and cortical thickness.

### Statistical Analysis

Student t tests were used to assess mean differences. Pearson correlation coefficient r values were calculated, multivariate regression analyses and mediation analyses were performed. IBM SPSS statistics version 24.0 was used for statistical analyses. P < 0.05 was considered statistically significant.

## RESULTS

### Group Comparisons

#### MS patients have increased cortical and white matter ‘glial activity load on PET (GALP)’ than healthy ***controls***

CoGM-lnGALP and WM-lnGALP scores were both increased in MS vs HC (10.0 ± 1.5 vs. 7.5 ± 1.4, +33% p=0.0003, Figure 1a; and 9.8 ± 1.5 vs. 6.6 ± 2.3, +48%, p=0.0002, Figure 1b, respectively). Meanwhile, CoT, normalized whole brain volume (nBPV), and normalized thalamic volume (ThV) were all decreased in MS vs HC (2.48 ± 0.07 vs 2.55 ± 0.08 mm, -3%, p=0.041, Figure 1c; 1404.3 ± 77.6 vs 1491.3 ± 51.7 ml, -6%, p=0.008, Figure 1d; and 20.1 ± 1.9 vs 22.8 ± 1.0 ml, -12%, p=0.001, Figure 2c, respectively).

**Figure 1.**
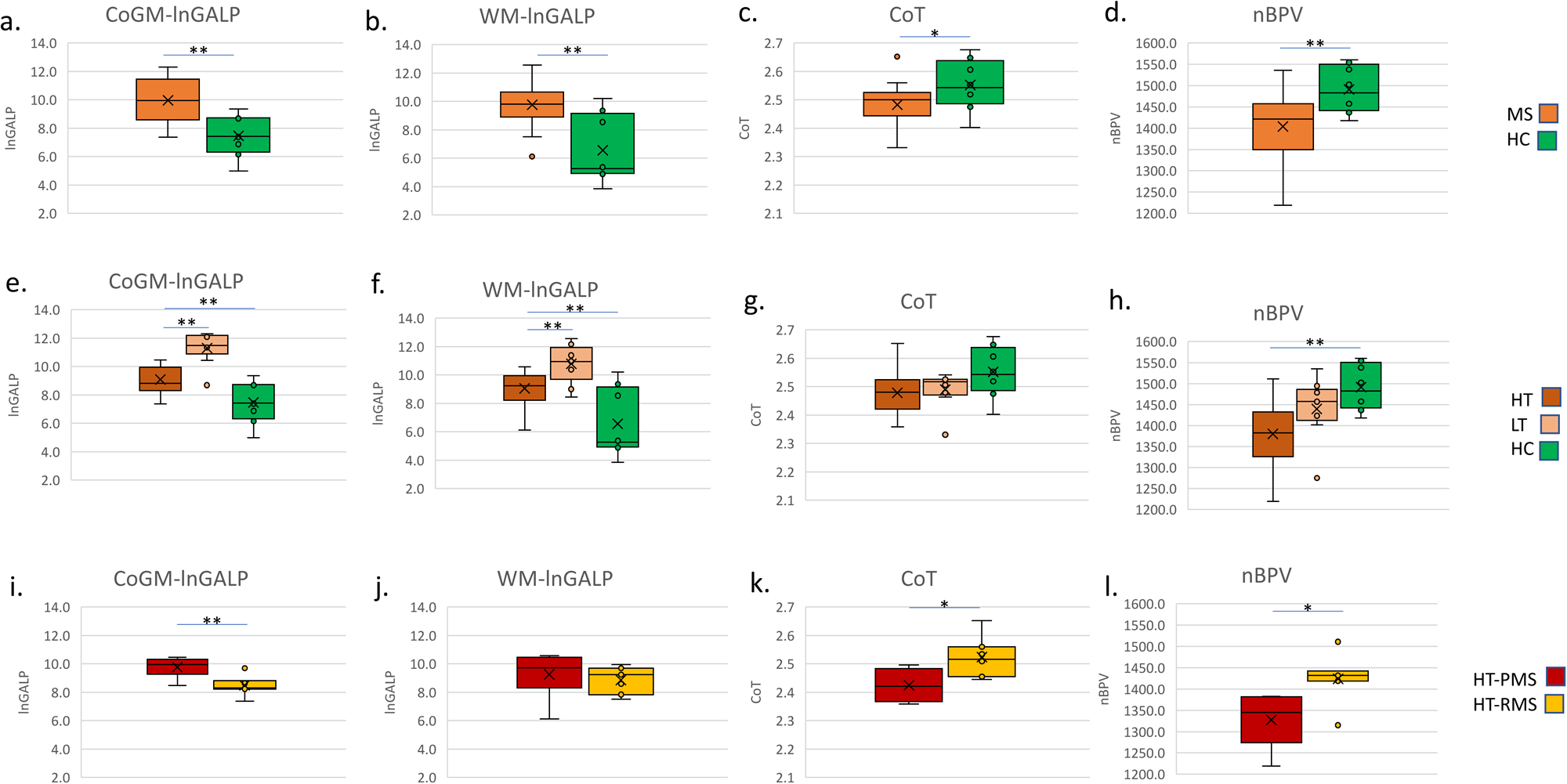
Group comparisons of lnGALP scores in the cortical grey matter and white matter, cortical thickness, and normalized brain volume in MS as compared to HC (a-d); in MS patients on high-efficacy DMT (HT) as compared to MS patients on low-efficacy or no DMT (LT) and as compared to HC (e-h); and in progressive vs relapsing MS patients in the HT group (i-l).

**Figure 2.**
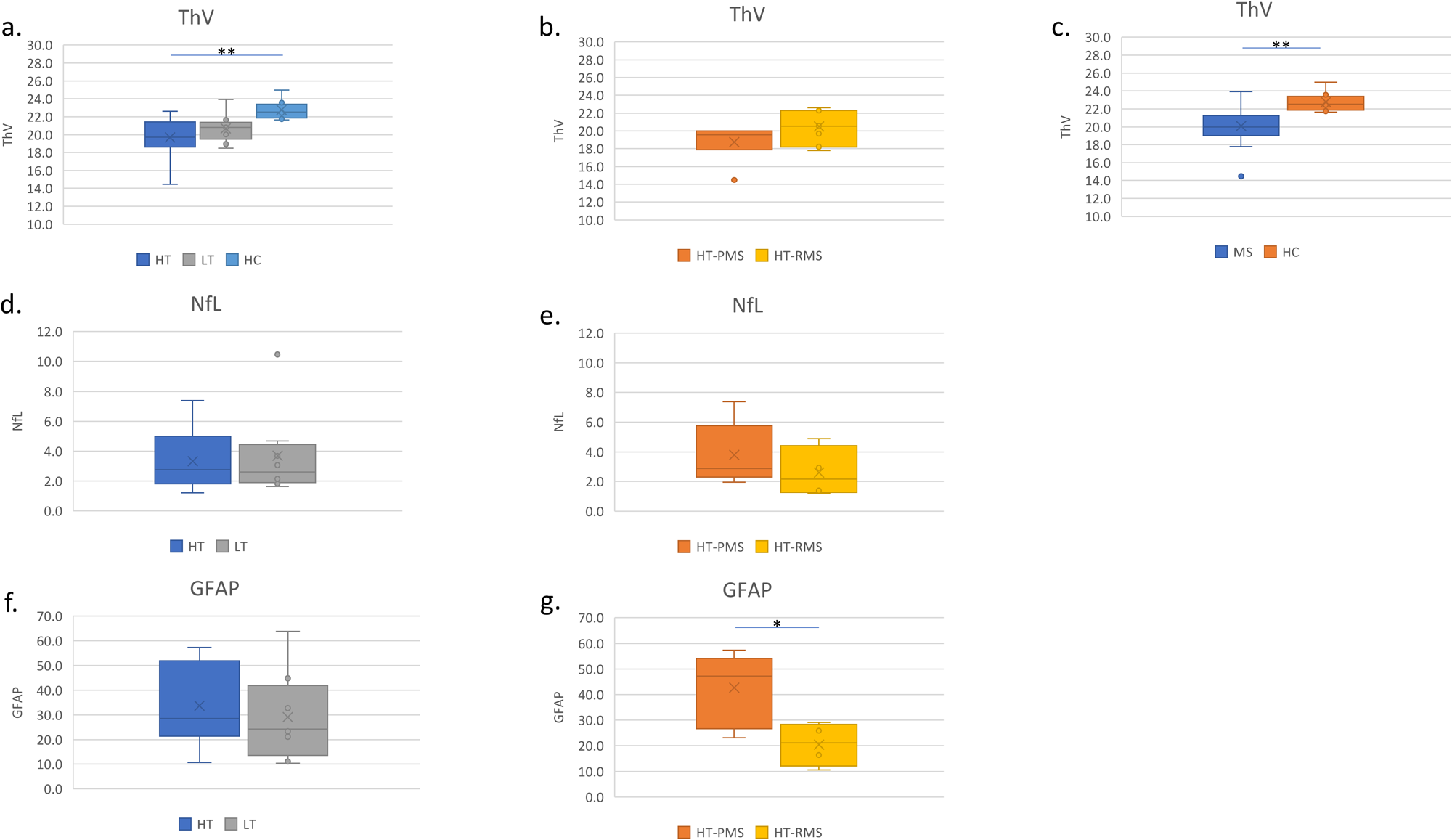
Group comparisons of thalamic volume in (a) MS patients on high-efficacy DMT (HT) vs MS patients on low-efficacy or no DMT (LT) vs HC; (b) in progressive MS patients on high-efficacy DMT (HT-PMS) vs relapsing MS patients on high-efficacy DMT (HT-RMS), and (c) in MS vs HC. (d-g) Group comparisons of NfL and GFAP in HT vs LT and in HT-PMS vs HT-RMS.

#### MS patients treated with high efficacy DMTs (HT-MS) have lower GALP than those treated with low efficacy DMTs (LT-MS) but are still abnormal as compared to HCs

CoGM-lnGALP and WM-lnGALP scores were both decreased in HT vs LT (9.1 ± 0.9 vs. 11.3 ± 1.1, -20%, p=0.00008, Figure 1e; and 9.1 ± 1.2 vs. 10.8 ± 1.3, -16%, p=0.006, Figure 1f, respectively). On the other hand, there were no significant differences in CoT, nBPV, ThV, sGFAP and sNfL between HT vs LT groups.

Mean CoGM-lnGALP and WM-lnGALP scores in the HT group of MS patients were both still increased compared to HC (9.1 ± 0.9 vs 7.5 ± 1.4, +21%, p=0.006, Figure 1e; and 9.1 ± 1.2 vs 6.6 ± 2.3, +37.8%, p=0.006, respectively, Figure 1f). While nBPV and ThV were decreased in HT vs HC (1379.8 ± 73.4 vs 1491.3 ± 55.3 ml, p=0.002, Figure 1h; and 19.7 ± 2.2 vs 22.8 ± 1.0 ml, p=0.002, Figure 2a, respectively), but the difference between mean CoT in HT vs HC did not attain statistical significance (2.48 ± 0.08 vs 2.55 ± 0.08 mm, p=0.068, Figure 2g).

#### Progressive MS patients have increased cortical glial activity as compared to relapsing-remitting MS patients despite treatment with high-efficacy DMTs

Within the HT-MS group, CoGM-lnGALP scores were increased in PMS vs RMS (9.8 ± 0.7 vs 8.5 ± 0.7, +15%, p=0.008, Figure 1i). However, WM-lnGALP was not significantly different in HT-PMS vs HT-RMS (9.3 ± 1.5 vs 8.9 ± 0.9, +4.5%, p=0.59). In terms of volumetric measures within the HT group, CoT and nBPV were both decreased in PMS vs RMS (2.43 ± 0.05 vs 2.52 ± 0.06 mm, -3.5%, p=0.017, Figure 1k; and 1327.7 ± 57.6 vs 1424.4 ± 53.4 ml, -6.8%, p=0.015, Figure 1l) but ThV was not statistically significant in HT-PMS vs HT-RMS (18.8 ± 1.9 vs 20.5 ± 1.9 ml, -8.3%, p=0.16, respectively). Interestingly, GFAP was increased in PMS vs RMS (42.61 ± 12.96 vs 20.50 ± 7.40, +108%, p=0.024, n=18, Figure 2g). However, within the same subset, NfL was not significantly different in PMS compared to RMS (3.78 ± 1.92 vs 2.60 ± 1.48, +45%, p=0.38, n=18).

### Correlations across groups

#### Increased cortical and white matter glial activation is linked with cortical atrophy

CoGM-lnGALP and WM-lnGALP scores were negatively correlated with CoT across MS and HC groups (r=-0.44 and r=-0.48, p<0.05, Figure 3a-b). On the other hand, there was a significant inverse relationship between ThV and WM-lnGALP (r=-0.404, p=0.027, Figure 3d) but Th-V only showed a trend towards an inverse relationship with CoGM-lnGALP (r=-0.310, p=0.096, Figure 3c). The relationship between nBPV and CoGM-lnGALP and WM-lnGALP did not achieve statistical significance.

**Figure 3.**
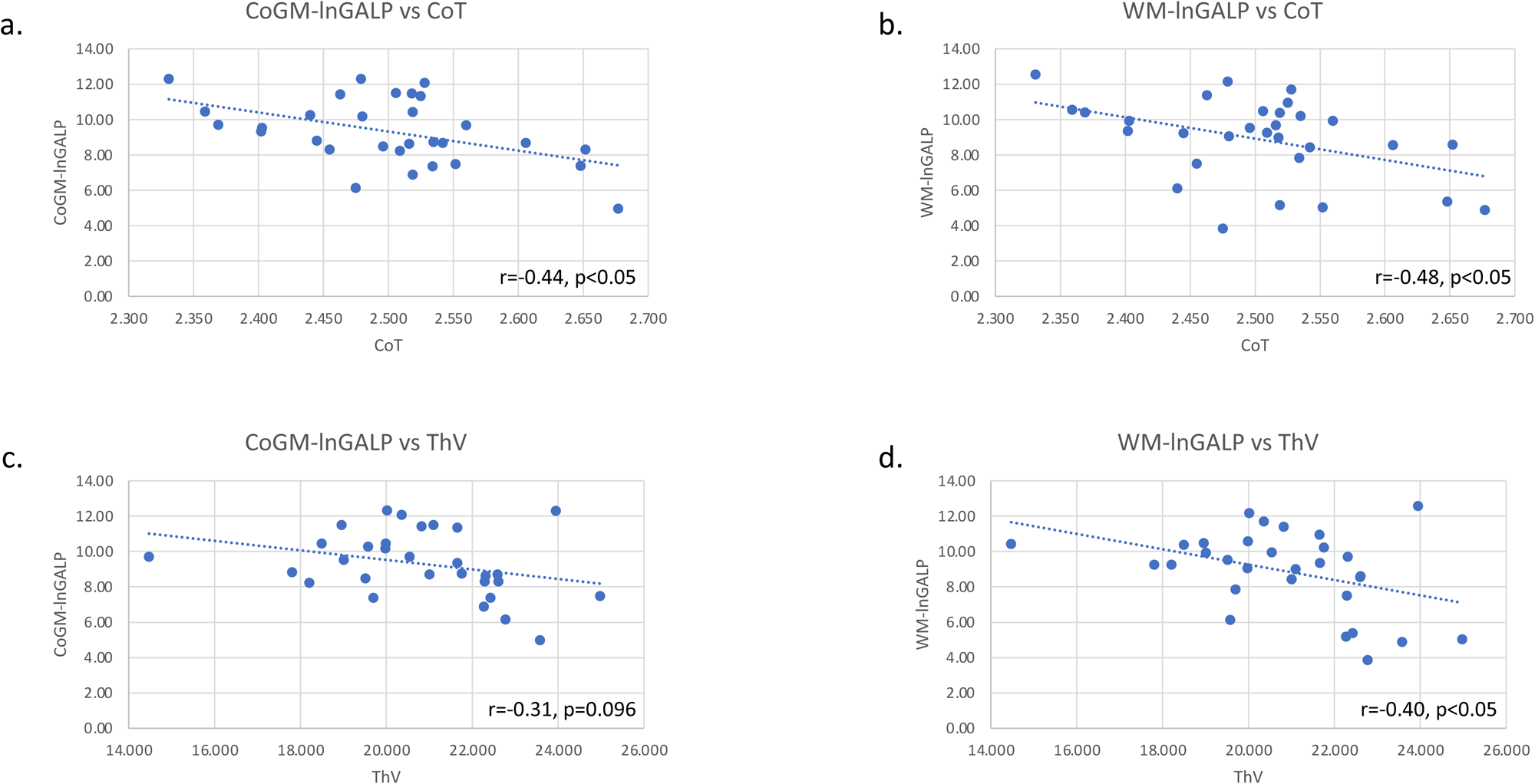
Correlations of lnGALP scores in the cortical grey matter and white matter with cortical thickness and normalized brain volume in MS and HC (a-d).

### Correlations within subgroups

#### Cortical glial activity is associated with clinical disability, fatigue scores, cortical degeneration and serum GFAP in HT-MS patients

On *subgroup analyses*, within the HT-MS group, CoGM-lnGALP correlated positively with EDSS, T25FW, and MFIS (r=0.645, 0.786, and 0.75, all p<0.05, Figure 4a-c). and negatively correlated with CoT (r=-0.659, p<0.05, Figure 4d) but did not correlate with nBPV or Th-V. CoGM-lnGALP was also positively correlated with sGFAP (r=0.67, p<0.05, Figure 4e). In contrast, there were no significant correlations between WM-lnGALP and the clinical (EDSS, T25FW, MFIS), MRI (CoT, nBPV, ThV) or serum biomarker measures in the HT group.

**Figure 4.**
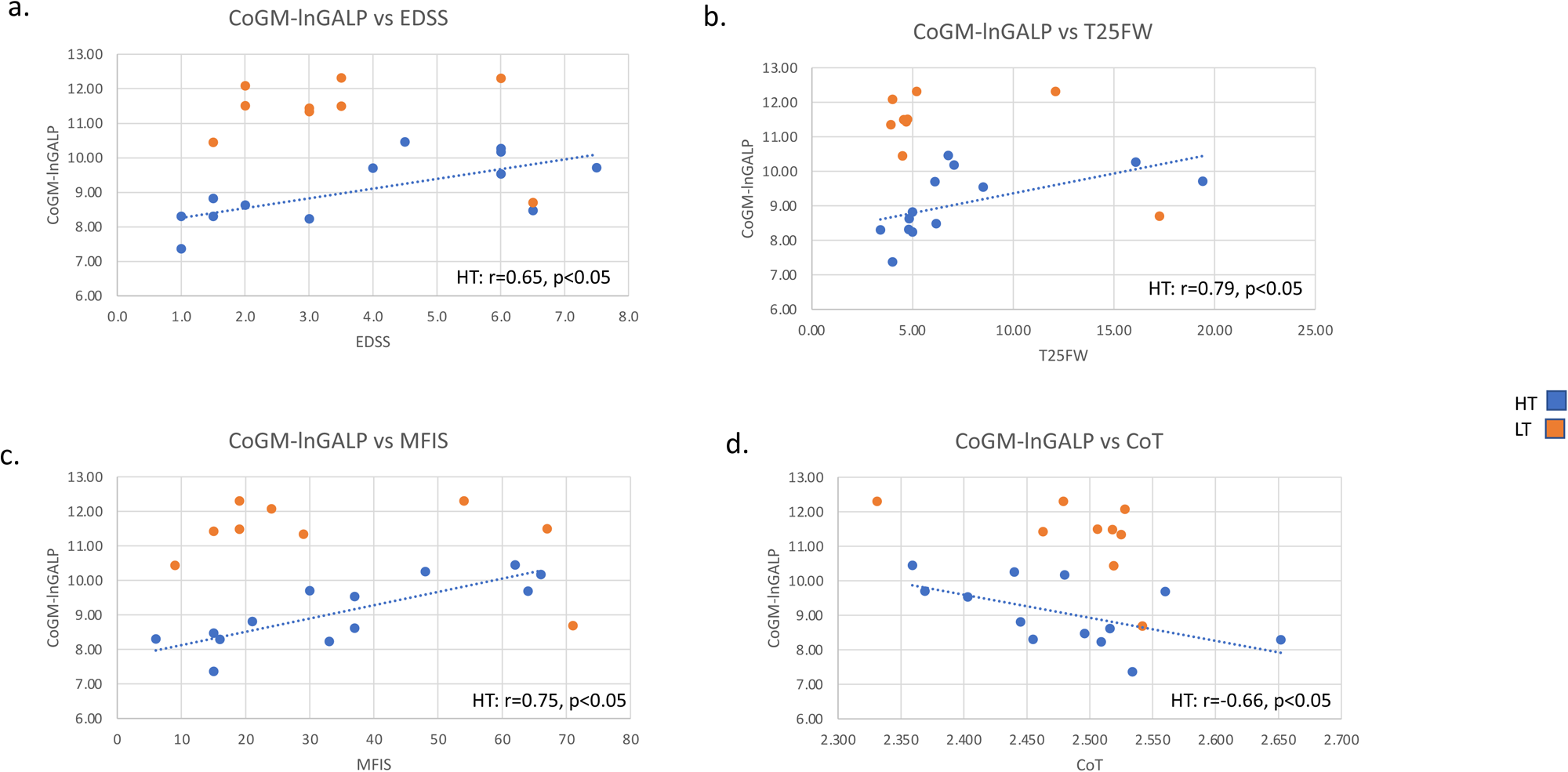
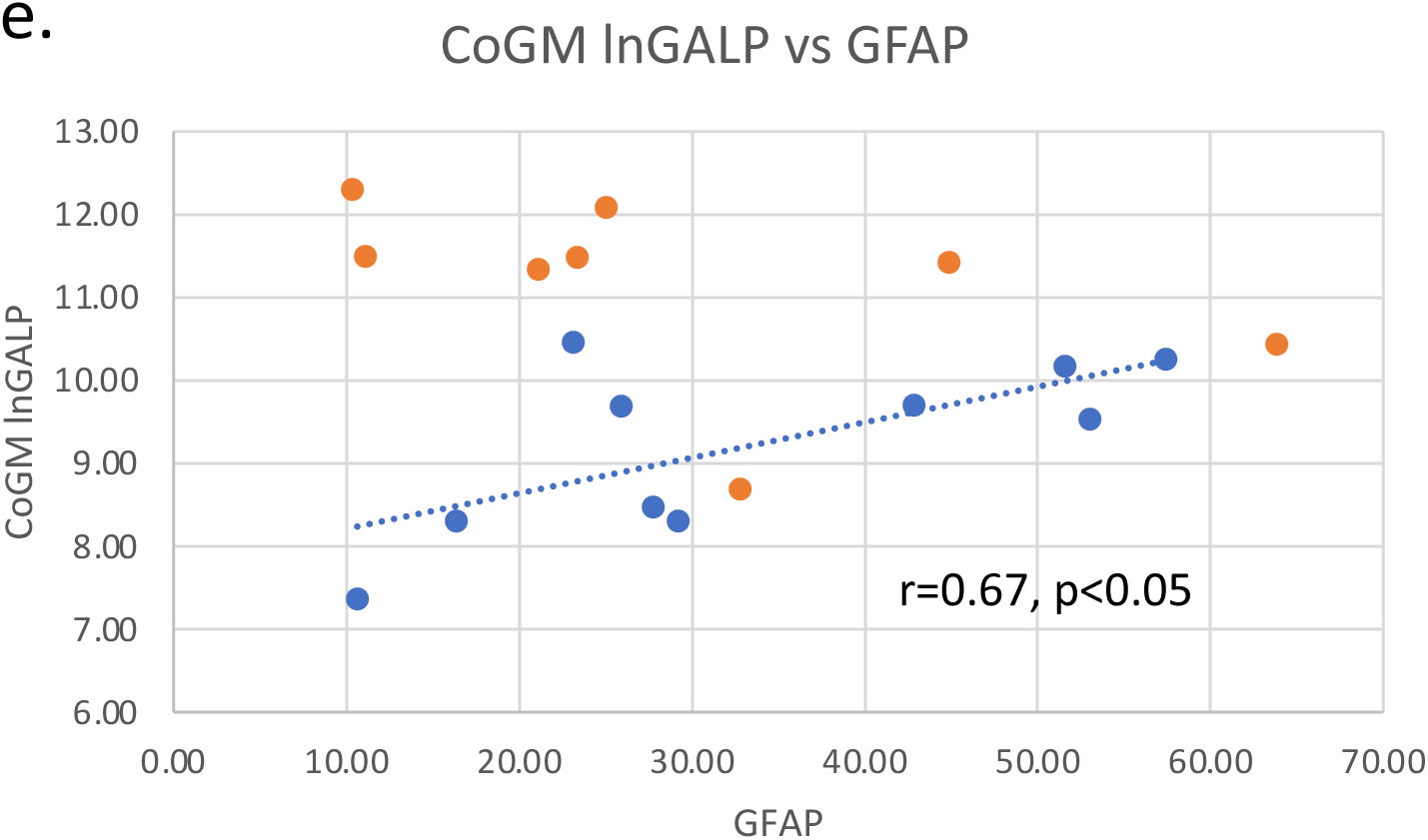
Correlations of lnGALP scores in the cortical grey matter with EDSS (a), T25-FW (b), MFIS (c), cortical thickness (d), and GFAP (e) in MS patients (HT=MS patients on high-efficacy DMT and LT= MS patients on low-efficacy or no DMT).

**Figure 5.**
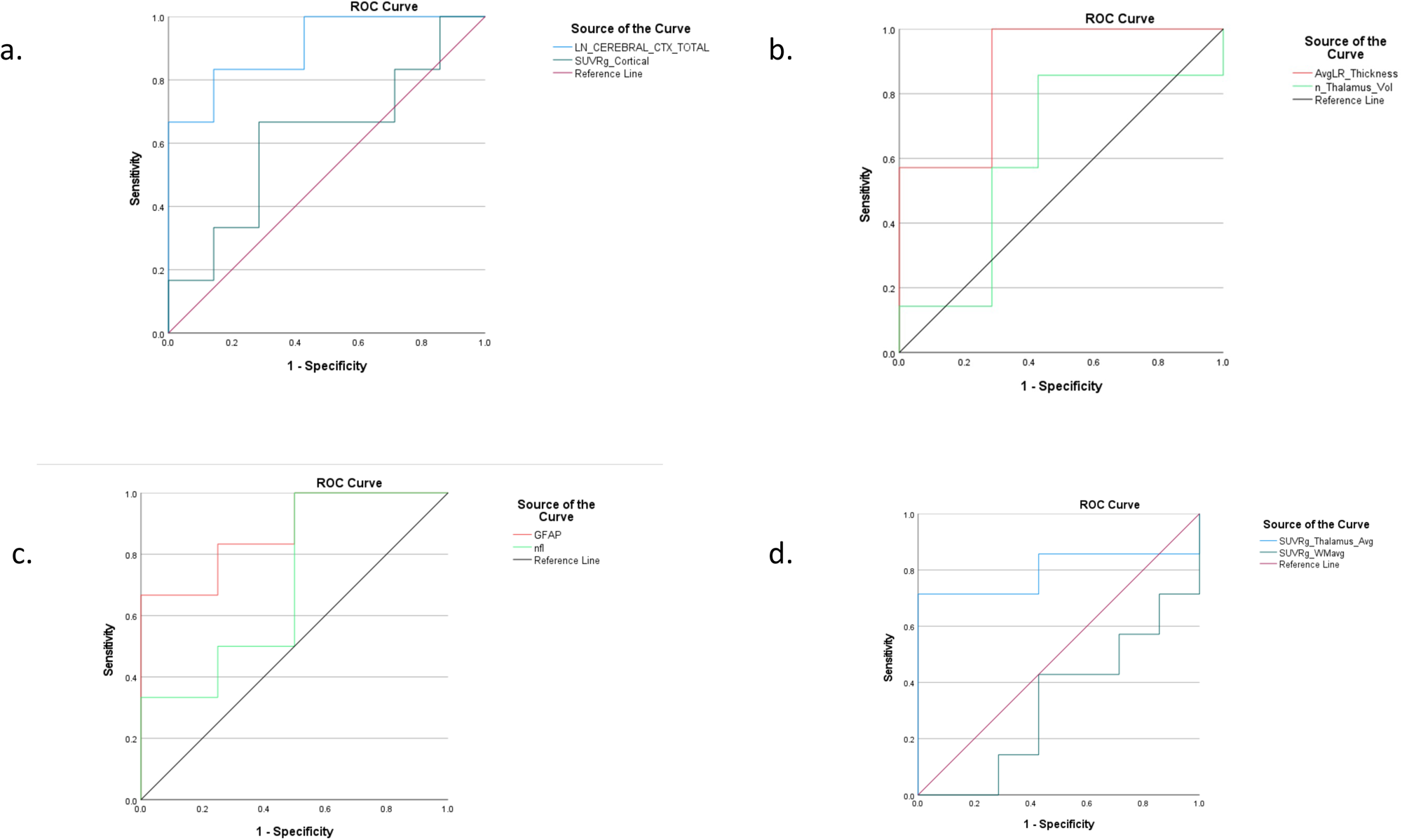
ROC analysis for classifying progressive MS vs relapsing-remitting MS by comparing lnGALP scores in the cortical grey matter vs cortical global SUVRs (a), cortical thickness vs thalamic volume (b), GFAP vs NfL (c), and global SUVRs of the thalamus vs white matter (d) in the HT group. (HT=MS patients on high-efficacy DMT)

**Figure 6a.**
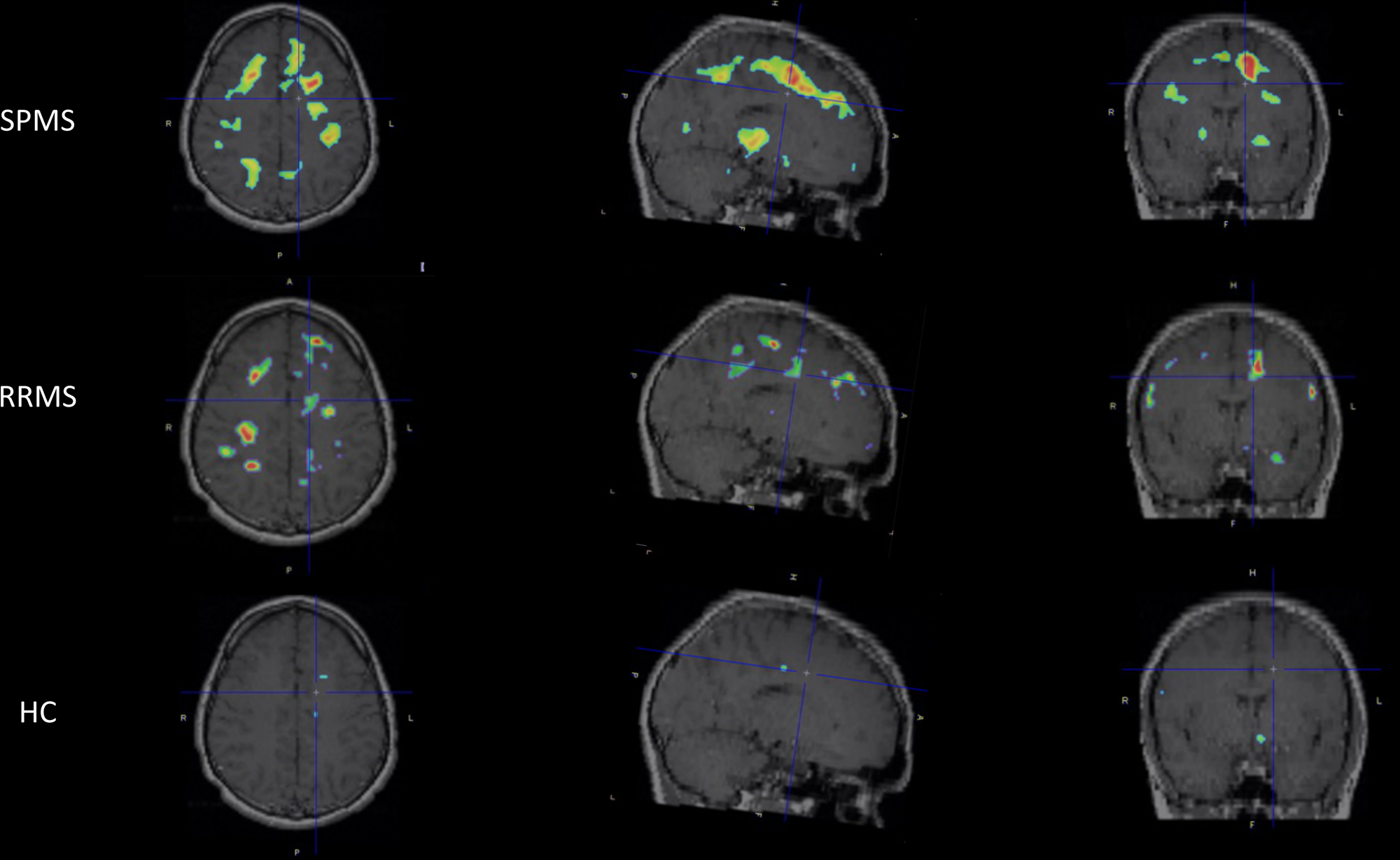
Individualized parametric z-score maps of the brain showing ‘Glial activity load on PET’ (GALP) in the brain, compared between a SPMS (top row), RRMS (middle row) and healthy control (bottom row) in transaxial, sagittal and coronal sections.

**Figure 6b.**
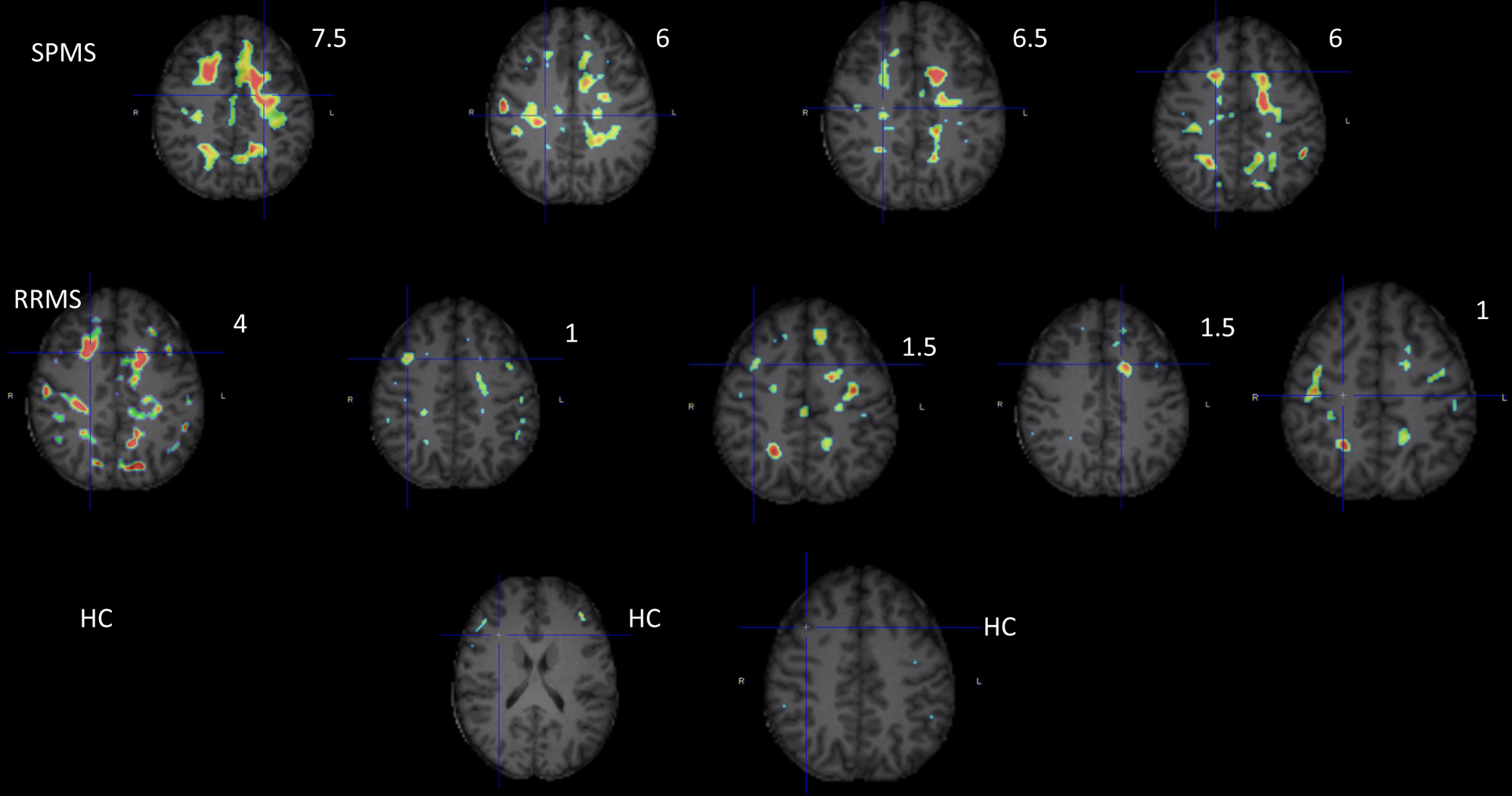
Transaxial sections of a subset of individual SPMS (top row, four SPMS subjects), RRMS (middle row, five SPMS subjects) and HC subjects (bottom row, 2 HC subjects) showing higher PET signal (GALP) in SPMS versus RRMS subjects and also demonstrating a relationship between with higher PET signal intensity and EDSS values (Numerical values on the top right corner of individual transaxial images refer to the EDSS score of each individual subject in the top and middle rows). EDSS=Expanded Disability Status Scale.

**Figure 7.**
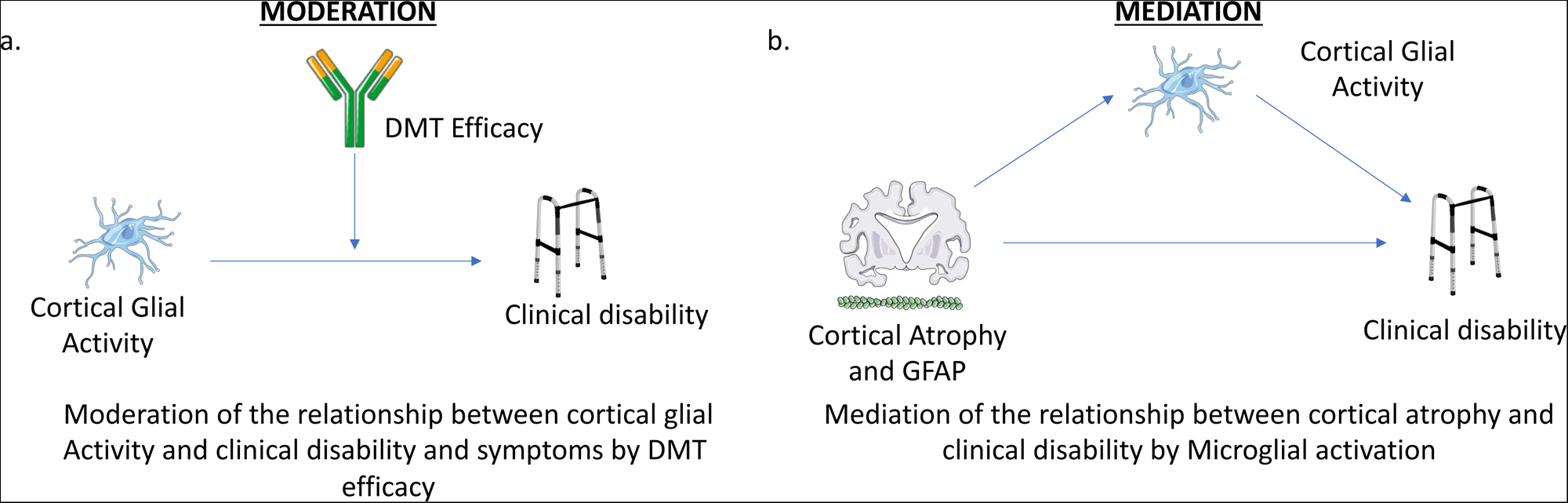
Schematic representation of a) moderation analysis shows the relationship between cortical glial activity and clinical disability is moderated by DMT efficacy and b) exploratory mediation analysis suggesting that the relationship between cortical atrophy/serum GFAP and clinical disability is mediated by cortical glial activity in HT-MS patients (HT=MS patients on high-efficacy DMT).

Within LT-MS group, Interestingly, none of the relationships between cortical and white matter GALP scores and clinical and morphometric scores in the LT group attained statistical significance. These findings emphasize the specificity of the relationship between cortical microglial activation with cortical thinning and clinical disability in the HT group.

#### DMT efficacy modulates the relationship between cortical glial activity and clinical impairment and serum biomarkers in MS patients

On multivariate analyses, CoGM-GALP predicted EDSS, T25FW, and MFIS scores, and GFAP and NfL levels independent of DMT efficacy but each of the relationships was moderated by the interaction between the DMT efficacy and cortical GALP. Further, CoGM-lnGALP predicted a diagnosis of progressive MS after adjusting for DMT efficacy, and this relationship was moderated by the interaction between the DMT efficacy and CoGM-GALP.

### ROC analysis

#### Cortical GALP outperforms cortical SUVR for differentiating progressive from relapsing-remitting patients in HT-MS group

On ROC analysis across modalities, CoGM-lnGALP, nBPV, CoT, and GFAP all provided significant differentiation for distinguishing progressive MS from relapsing MS within the HT group (AUCs= 0.91, 0.90, 0.88, and 0.88, respectively). Meanwhile, NfL, WM-lnGALP, and ThV had lower AUCs on ROC analysis (0.71, 0.67, and 0.61, respectively). On further ROC analysis across additional PET indices, SUVg, SUVTh, and SUVRgTh were also significant in distinguishing progressive MS from RMS within HT (AUC= 0.81, 0.81, and 0.76, respectively). SUVRgCo and WM SUV had lower AUCs within HT (0.62 and 0.60, respectively).

### Mediation analysis

#### Cortical neuroinflammation mediates the relationship between progressive MS and cortical atrophy and serum GFAP levels

While both lower cortical thickness and higher serum GFAP predicted a diagnosis of progressive MS in the HT-MS group, this relationship was mediated by cortical glial activity (CoGM-GALP) (Sobel’s test p-value <0.05) on exploratory analyses, supporting the central role of glial activation in pathogenesis of progressive MS.

## DISCUSSION

There are several key findings in our paper. Firstly, we describe a novel, unbiased, individualized approach to analyzing [F-18]PBR06-PET in MS. Secondly, we demonstrate that in MS patients, high efficacy disease modifying therapies may reduce but do not normalize glial activity in the cerebral cortex and white matter. Further, we demonstrate that among MS patients treated with high efficacy DMTs, cortical smoldering inflammation measured by [F-18]PBR06-PET is linked with cortical atrophy (lower cortical thickness), worsening physical disability, higher depression and fatigue scores, higher serum GFAP levels, and is increased in progressive MS as compared to relapsing MS,.

Voxel-wise, z-score mapping (VZM) has been extensively applied in brain PET imaging and recommended for enhanced evaluation of individual patients’ FDG-PET scans.^23^ This approach has led to a standardized, unbiased approach for interpreting brain FDG-PET images in the evaluation of dementias.^24^ It has also yielded high sensitivity and specificity in distinguishing patients with Alzheimer’s disease from patients with other dementias and from healthy controls.^24^ A key advantage of the VZM approach is that it can be applied to static PET scans acquired at later time points following radiotracer injection when the radiotracer is more likely to have reached an equilibrium state. This approach is widely used clinically because it is less time consuming as compared to dynamic imaging and improves patient comfort. From an image quality standpoint, [F-18]PBR06 is a good candidate radiotracer for the application of the VZM approach on static imaging as it is a [F-18] labeled PET tracer with a longer (∼110 minutes) half-life. Owing to its longer half-life, static imaging acquired 60-90 minutes following radiotracer injection provides an improved signal-to-noise ratio and better image quality as compared to [C-11]-labeled tracers, which have a significantly shorter half-life (20 minutes) and therefore result in poor count statistics at later time points.^12^

There is an urgent need to standardize image analysis and interpretation in TSPO-PET imaging. We have found that individualized, VZM mapping is feasible and provides meaningful results in MS patients using [F-18]PBR06. Previous methods in TSPO-PET have focused on evaluating innate immune activation based on average estimates of radiotracer uptake in large brain regions of interest (ROIs). However, smaller, focal areas of the brain parenchyma may have abnormalities that may be missed using the ROI-approach, reducing its sensitivity. Furthermore, using a uniform threshold cut-off to define abnormalities for the entire brain may not be accurate due to the known regional variations in microglial density in healthy brains. On the other hand, a voxel-wise, z-score approach identifies specific abnormalities in a given voxel in a patient’s image as compared to the corresponding set of voxels in the reference population, thereby improving the sensitivity and specificity of the results. Pre-defined smoothing parameters and z-score thresholds are crucial to minimizing the impact of image noise and false-positives due to multiple statistical comparisons in VZM. In our study, we applied a 3-dimensional smoothing filter during image processing and a z-score threshold of >4 to address these issues.

We have previously reported our preliminary findings using the standardized uptake value ratio (SUVR) approach in a subset of the current sample that showed an inverse relationship between thalamic PET ligand uptake and brain volume.^5^ Using the VZM approach, we are able to demonstrate an inverse relationship between innate immune activation in the cerebral cortex and white matter and thalamic and whole brain volumes. These results reinforce the relationship between innate immune activation and brain parenchymal tissue loss in a larger dataset, and using our new, individualized, VZM methodology.

We demonstrate that patients treated with high efficacy DMTs have lower innate immune activation than those who are either untreated or treated with low efficacy disease modifying therapies. Several DMTs have been previously shown to reduce PET microglial activation in MS (largely, RRMS) patients in longitudinal studies. A study in a mixed population of RRMS and SPMS patients showed a reduction in [C-11]PK11195-PET distribution volume ratios after one year of treatment with natalizumab.^25^ Similarly, 12% reduction in lesional BPnd was seen after fingolimod treatment for 6 months in a RMS population.^26^ Further, our preliminary analysis showed a reduction of 19.4% in cortical glial activity load on PET (‘GALP”) after 3 months of treatment with Ofatumumab, a B-cell therapy.^13^ On the other hand, although it is not straightforward to compare across studies, it is notable that treatment with glatiramer acetate showed only a 3.17% reduction in microglial activation in RMS patients after 1 year.^27^ Overall, these longitudinal studies suggest that higher efficacy DMTs are associated with significantly reduced microglial activation in at least some brain regions. Our data in this current cross-sectional study is consistent with these longitudinal studies in terms of decreased PET signal in HT vs LT group. Moreover, the magnitude of longitudinal change following initiation of a high-efficacy treatment in these studies is similar to the cross-sectional group differences between the HT and LT groups (∼20%) we report in our study.

Further, it is well known that the currently approved DMTs have not shown efficacy in inactive progressive MS patients and a significant proportion of patients progress despite being on a high efficacy DMT.^28, 29^ This phenomenon may be explained by our finding that although high efficacy DMTs are associated with a decreased PET signal, there is still abnormally persistent glial density load (or smoldering inflammation) despite high efficacy treatment in MS patients. Many of the current high efficacy DMTs target the peripheral immune system, do not cross the blood brain barrier and therefore have limited efficacy on the progressive component of the disease process.^29, 30^ We believe that our approach can enrich clinical trials by identification of patients with significant “residual” microglial activation despite high-efficacy DMTs who may benefit from emerging treatments targeting smoldering inflammation. We also believe that the GALP approach can facilitate individualized assessments of treatment response in MS patients.

Cortical microglial activation and neurodegeneration have been associated with MS progression in multiple pathological and imaging studies.^31, 32^ Within the high efficacy DMT group, we observed higher abnormal cortical microglial activation in SPMS as compared to RMS patients. This is consistent with increased regional cortical TSPO-PET signal, which we have reported previously using [F-18]PBR06 and has also been reported using [C-11]PK11195.^5, 33^ Further, we observed an inverse relationship between cortical GALP scores and cortical thickness. Cortical atrophy is a key feature of MS pathology and cortical microglial activation associated with synaptic injury has been consistently reported in pathological studies.^31, 32, 34^ Our findings provide novel data reinforcing the link between abnormal innate immune activation and local brain parenchymal injury in the cortical grey matter in MS patients despite high efficacy treatment. Furthermore, we observed that abnormal cortical GALP scores predicted clinical disability, depression and fatigue severity, highlighting the clinical relevance and validity of cortical microglial activation and our methodology in these MS patients. Prospective longitudinal studies in MS patients are urgently needed to investigate the prognostic value of abnormal cortical microglial activation detected by [F-18]PBR06-PET. Serum GFAP is an emerging biomarker for progressive MS^35^ and interestingly, our findings suggest that cortical innate immune activation is linked with elevated serum GFAP levels in patients treated with high-efficacy MS DMTs. GFAP is an astrocytic intermediate filament and astrocytes are known to contribute to the TSPO-PET signal in MS.^35^ The relationship between TSPO-PET and serum GFAP warrants further detailed evaluation.

There are several limitations to our study. Our sample size is small and further validation of our results in future larger studies is needed. From a technical standpoint, voxel-wise methods may be prone to noisy estimation of parameters and assessment of test-retest variability is needed, although our preliminary data for evaluating longitudinal changes after therapy are encouraging. Refinement of image processing steps may improve sensitivity for detection of abnormal regional microglial activation.

In summary, we have reported a novel voxel-wise, z-score based approach to quantitate abnormal innate immune activation using [F-18]PBR06-PET in MS. This study provides PET evidence for clinical and patho-biological importance of this ‘smoldering’ inflammation that persists in MS patients despite treatment with high efficacy disease modifying therapies. This PET approach is clinically feasible, is relevant for multicentric studies, and can provide meaningful insights into MS pathology and response to treatment in individual patients. Further, it can potentially aid evaluation of progression independent of relapse activity (PIRA) and expedite therapeutics research in this area, which is urgently needed to address the unmet need of targeting innate immune activation/smoldering inflammation in MS.

## Data Availability

All data produced in the present study are available upon reasonable request to the authors.

## Notes

### Competing Interest Statement

Tarun Singhal has received research support from Novartis Pharmaceuticals and Genzyme-Sanofi, consulting fees from Novartis Pharmaceuticals, Genentech, and EMD Serono, speaking fees from Tiziana Life Sciences, and research funding
from Nancy Davis Foundation's "Race to Erase MS" program, Ann Romney Center for Neurologic Diseases, Harvard Neuro-Discovery Center, National Multiple Sclerosis Society, Department of Defense, and Water Cove Charitable Foundation.
Eero Rissanen has received a fellowship grant from the Sigrid Juselius Foundation and research grant from the Sakari Alhopuro Foundation.
Tanuja Chitnis has received compensation for consulting from Biogen, Novartis Pharmaceuticals, Roche Genentech and Sanofi Genzyme, and has received research support from the National Institutes of Health, National MS Society, US Department of Defense, Sumaira Foundation, Brainstorm Cell Therapeutics, EMD Serono, I-MAB Biopharma, Mallinckrodt ARD, Novartis Pharmaceuticals, Octave Bioscience, Roche Genentech and Tiziana Life Sciences.
Rohit Bakshi has received consulting fees from Bristol-Myers Squibb and EMD Serono and research support from Bristol-Myers Squibb, EMD Serono and Novartis.
Howard L. Weiner has received research support from Cure Alzheimer's Fund, EMD Serono Inc., Genentech Inc., National Institutes of Health, National Multiple Sclerosis Society, Sanofi Genzyme, and Verily Life Sciences, and has received payment for consulting from Genentech, Inc, IM Therapeutics, I-MAB Biopharma, MedDay Pharmaceuticals, Tiziana Life Sciences and vTv Therapeutics.

### Clinical Trial

NCT02649985

### Funding Statement

This study was funded by Nancy Davis Foundation's "Race to Erase MS" program, Ann Romney Center for Neurologic Diseases, Harvard Neuro-Discovery Center, and Water Cove Charitable Foundation.

### Author Declarations

IRB of Mass General Brigham gave ethical approval for this work.

