## Supplementary Tables for "Glial activity load on PET (GALP) reveals persistent ‘smoldering’ inflammation in MS despite disease modifying treatment: [F-18]PBR06 study"

Supplementary Table 1

ROC analysis within HT group for classifying progressive MS

AUC across modalities for classifying progressive MS

| Modality | AUC |
| --- | --- |
| CoGM-lnGALP | 0.91 |
| WM-lnGALP | 0.67 |
| CoT | 0.88 |
| BPV | 0.898 |
| Th-V | 0.612 |
| GFAP (n=10) | 0.88 |
| NFL (n=10) | 0.71 |

Supplementary Table 2

AUC across PET indices for classifying progressive MS

| PET index | AUC |
| --- | --- |
| CoGM-lnGALP | 0.905 |
| WM-lnGALP | 0.67 |
| SUVRgTh | 0.762 |
| SUVRgCo | 0.62 |
| Global SUV | 0.81 |
| WM SUV | 0.595 |
| Th SUV | 0.81 |

Supplementary Table 3.

Correlation coefficients for comparisons of clinical measures and MRI with other PET indices in HT group

|  | EDSS | T25FW | CoT | Th-V | nBPV |
| --- | --- | --- | --- | --- | --- |
| CoGM-lnGALP | 0.645 (0.017)* | 0.786 (0.001)** | -0.659 (0.014)* | -0.115 (0.707) | -0.280 (0.354) |
| WM-lnGALP | 0.452 (0.121) | 0.401 (0.174) | -0.302 (0.316) | -0.198 (0.517) | -0.357 (0.231) |
| SUVRgTh | 0.601 (0.030)* | 0.440 (0.113) | -0.341 (0.255) | -0.297 (0.325) | -0.549 (0.052) |
| SUVRgCo | -0.064 (0.836) | -0.038 (0.901) | -0.209 (0.494) | 0.357 (0.231) | 0.412 (0.162) |
| Global SUV PVC | 0.404 (0.170) | 0.434 (0.138) | -0.753 (0.003)** | -0.379 (0.201) | -0.434 (0.138) |
| WM SUV PVC | -0.044 (0.886) | -0.011 (0.972) | -0.308 (0.306) | -0.077 (0.803) | -0.093 (0.762) |
| Th SUV PVC | 0.391 (0.187) | 0.368 (0.216) | -0.692 (0.009)** | -0.385 (0.194) | -0.462 (0.112) |

* equals p<0.05; ** equals p<0.01
